# Developing a Health Score and Predicting disease Risks Using DKABio-clusters

**DOI:** 10.1101/2024.06.16.24308995

**Authors:** Kuang Fu Cheng, Ya-Hui Yang, Chih-Hsiung Su, Meng-Chun Tsai

## Abstract

In our research, accurately estimating the morbidity of individuals with specific conditions, plays a pivotal role in enhancing healthcare delivery systems. Introducing DKABio-clusters, we delve into their distinct characteristics, showcasing their profound implications for healthcare management. A primary focus of DKABio-clusters lies in developing a unique health assessment tool, termed DKABio-HS, alongside predictive risk analysis.

DKABio-HS facilitates the computation of a comprehensive “disease-related” score, condensing an individual’s health status into a singular numerical value. Our investigation reveals the remarkable consistency of this health score, with minimal variations observed between training and validation datasets (mean absolute percentage errors within 0 to 10 years remaining below 0.1%, with all mean absolute percentage errors ranging between 1.2-1.6%). A higher health score denotes better health or reduced disease risk, diminishing with age or the presence of multiple diseases.

Utilizing this health score, we establish a classification framework termed the “disease map,” enabling precise differentiation of individuals across various health states. Through this framework, individuals without diseases can be categorized as either healthy or sub-healthy, facilitating tailored health management strategies for preventive interventions. Our analysis indicates that individuals classified as sub-healthy exhibit significantly elevated disease risks compared to those deemed healthy (Female (male) 5-year risks of developing at least one disease are 29% vs. 15% (29% vs. 16.5%)).

Furthermore, leveraging a carefully selected set of health variables, we can delineate the distribution of DKABio-clusters and concurrently predict the 10-year risks associated with 15 diseases/conditions. Validating the predictive capabilities of our model, we compare predicted risks with true risks derived from extensive datasets, demonstrating non-statistically significant differences in the majority of cases. All analyses are grounded in data sourced from the National Health Insurance Research Database (more than 2 million participants) released by the National Health Research Institute, Taiwan and the Mei Jau Health Management Institution database (more than 0.75 million participants), spanning the years 2000 to 2016 in Taiwan.

## 1. Introduction

Emerging as a prominent field, precision health aims to proactively prevent diseases by harnessing cutting-edge technological advancements, data science, and artificial intelligence. In this context, we present a comprehensive approach that caters to individualized prevention and treatment, ensuring optimal well-being. Our pivotal step involves utilizing the DKABio (Data Knowledge in Action)-clusters to generate a “disease-related” health score (DKABio-HS) that condenses an individual’s health status into a single numerical value. Additionally, we leverage this score to predict the risks of 15 common chronic diseases or symptoms.

The American Thoracic Society defines health status as an individual’s relative level of wellness and illness, encompassing biological or physiological dysfunction, symptoms, and functional impairment. Accurately measuring health status plays a vital role in evaluating successful aging or active aging, among other factors. Successful aging indicators, including brisk walking, independence, emotional vitality, and self-rated health, have been correlated with mortality (Mount et al. [1]). Furthermore, the number of successful aging indicators exhibits strong associations with age and the Charlson Comorbidity Index. Lee et al. [2] employed exploratory factor analysis to establish a five-determinant model (comprising physical activity, life satisfaction and financial status, health status, stress, and cognitive function) to assess meaningful and successful aging indicators. Notably, health status emerged as the most influential factor in living independently and a crucial predictor of self-rated health. Similar factors have been previously linked to frailty (Lin et al. [3]).

Various health status measures have been developed for diverse purposes. For instance, the Elixhauser Index (Elixhauser et al. [4]) was devised using diagnoses reported in hospital discharge records, while chronic disease scores were introduced by Von Korff et al. [5] and Iommi et al. [6] based on prescription data. Additionally, Li et al. [7] developed polygenic risk scores for disease risk prediction. Pano, et al. [8] created health score for lifestyle and well-being index. In contrast, our health score, the DKABio-HS, serves unique objectives. It draws upon the DKABio clustering system, created using national insurance data and health examination data. The insights provided by the DKABio-HS and the subsequent risk predictions prove invaluable in formulating healthcare strategies for individuals in different health categories, such as the healthy, sub-healthy, and diseased populations. These aspects form the core tenets of precision health.

## 2. Methods

### 2.1 Participants and study design

Although the DKABio-HS and the Frailty Index (FI) share some similarities in their fundamental concepts, they also exhibit notable differences. The primary objective of the FI was to predict mortality risk and explore its factor structure, as evident in studies like the Taiwan FI (TwFI) conducted by Lin et al. [3]. Conversely, the DKABio-HS was primarily developed for disease control and health management in the context of precision health. While the TwFI proves valuable in aging research, the DKABio AI-HS finds its utmost utility in precision health management.

Both the TwFI and the DKABio-HS rely on similar data variables, including demographic information, subjective health evaluations, family and personal disease history, social behavior, and laboratory markers such as urine and blood tests. The original TwFI was derived from the SEABS (Social Environment and Biomarkers of Aging Study) dataset (Cornman et al. [9] (2016)), comprising 139 health-related variables collected from 1,284 participants aged 53 and above. However, a shorter version of the TwFI, based on only 35 health variables, demonstrates properties compatible with the original TwFI. In contrast, the computation of the DKABio-HS utilized 148 health variables primarily gathered by the Taiwan Mei Jau Health Management Institution from approximately 750,000 participants aged 20 and above between 2000, January 1 and 2016, December 31 (referred to as Data A). The average observation period per participant was approximately 3.6 years. Only a few health variables, such as cancer marker indexes, were obtained from cancer studies conducted by a hospital in central Taiwan and from questionnaires. For more detailed information on Data A, refer to Wu et al. [10].

It is important to note that the computation of the TwFI is relatively straightforward, involving the ratio of the summed health deficits scores to the total health deficits items. In contrast, the DKABio-HS employs powerful machine learning techniques, namely hierarchical clustering analysis and logistic regression, to develop the fundamental structure of the score. This structure is crucial for generating risk predictions as well.

This study was a retrospective analysis of medical records. All data were collected in compliance with Taiwan’s “General Data Protection Regulation” and were fully anonymized before being accessed by the authors. The study received approval from the Ethical Review Committee of National Taiwan University (NTU-REC No.: 202402EM002) for the use of Data A and Data B (detailed below), which the authors began acquiring on March 6, 2024.

### 2.2 Computation Models

The computation of the DKABio-HS involves two main steps. The first step utilizes a hierarchical clustering algorithm, also known as unsupervised classification, with the Euclidean metric. This algorithm relies on comorbidity scores, age indexes, and gender to partition diseased participants into three distinct clusters. The comorbidity score, a variant of the Charlson Comorbidity Index, is based on 15 chronic diseases and conditions (refer to Table 1). The original Charlson Comorbidity Index, developed by Charlson et al. [12], is a weighted index used to predict the one-year risk of death for patients with specific comorbid conditions upon hospitalization. Deyo et al. [14] and Romano et al. [15] adapted the index to ICD-9-CM diagnosis and procedure codes and CPT-4 codes, respectively, enabling its calculation using administrative data. In our case, the weights for the comorbidity score were determined by rounding off the coefficients obtained from a regression model that utilized “out-patient dot” (money equivalent paid to healthcare service providers from the National Health Insurance Administration) as the response variable and 15 disease statuses as explanatory variables. The age indexes are calculated as

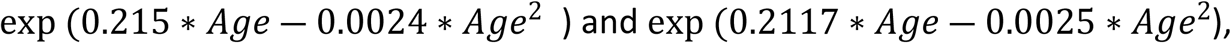

for females and males, respectively. For non-diseased participants, a similar clustering algorithm is applied to the continuous data, taking into account the out-patient dot, age indexes, and gender, resulting in their grouping into three different clusters as well. These clusters are referred to as DKABio-clusters. Table 1 summarizes the 10-year risks of 15 diseases/conditions for individuals belonging to each cluster. These cluster characteristics were derived from the National Health Insurance Research Database, released by the National Health Research Institute, Taiwan (refer to studies like Lin et al. [16] or Hsieh et al. [17]), which is known as Data B. The data were collected between 1997, January 1 and 2012, December 31 from 2 million participants of any age.

**Table 1.**
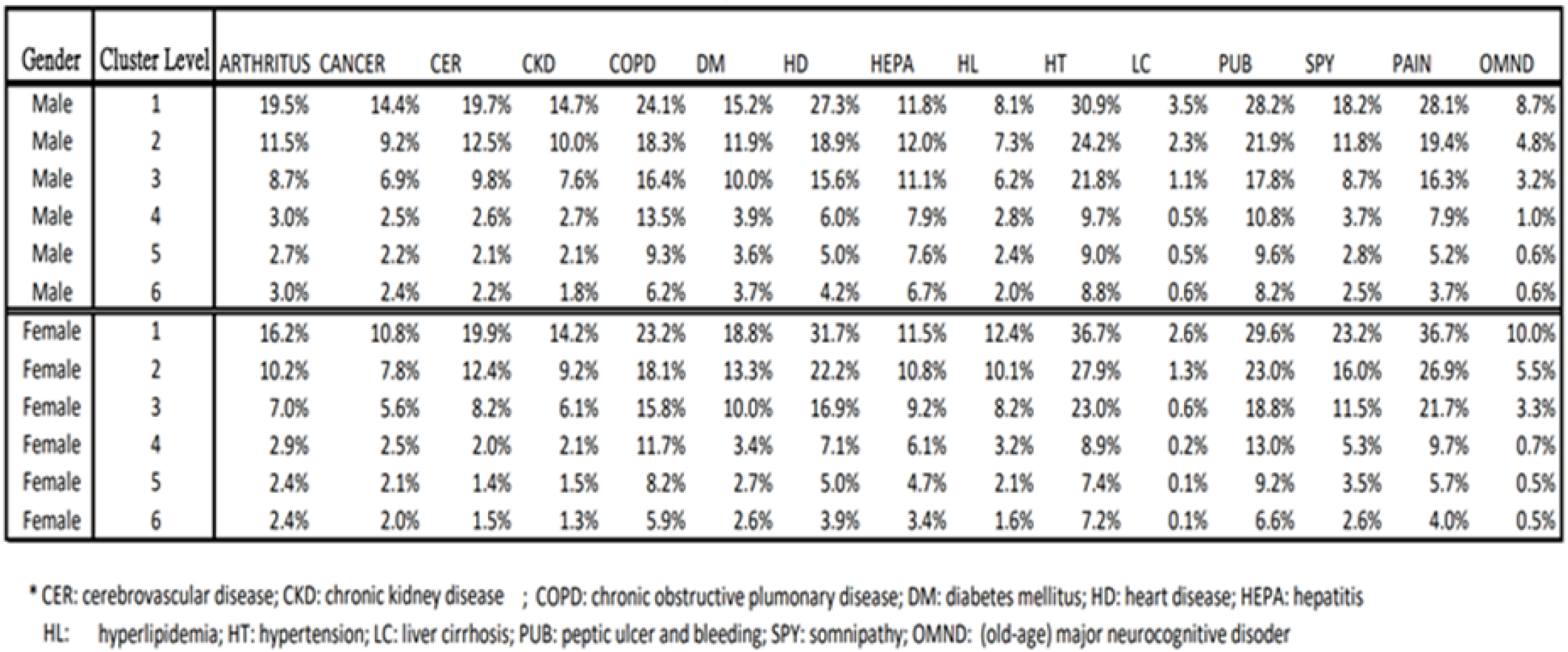
10-year Risks of diseases/sympotoms*.

The results presented in Table 1 demonstrate that the risks of all diseases or major symptoms generally decrease as the cluster level increases. Notably, for male (female) participants, the 10-year risk ratios of cluster level 6 compared to level 1 are greater than 5 for 9 (12) out of 15 diseases/symptoms. This suggests that the DKABio-cluster level (CL) variable is a potent risk predictor for many significant chronic diseases and symptoms.

The second step in calculating the HS involves estimating the distribution of the CL variable given specific values of health variables. This is accomplished by determining the transition probability *Pi* from cluster (state) *i* to cluster *i* ― 1, where *Pi* is calculated as follows:

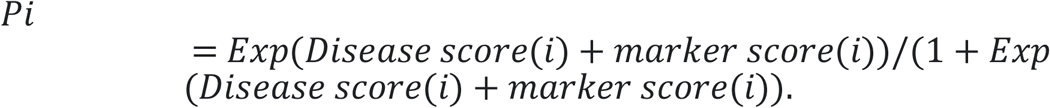

To elaborate on the general calculation of transition probability, let *Z*1 represent the number of reported diseases by a participant from hypertension, hyperlipidemia, diabetes mellitus, arthritis, chronic kidney disease, hepatitis, peptic ulcer, and bleeding. Similarly, *Z*_2_ represents the number of reported diseases from cerebrovascular disease, heart disease, chronic obstructive pulmonary disease, liver cirrhosis, cancer, somnipathy, (old-age) major neurocognitive disorder, and pain. The disease score is computed as

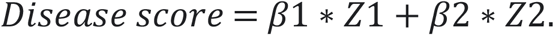

The participant’s observed health variables are denoted as *X*_*k*_, *k* = 1,..,*K*. For each disease *D*_*j*_ mentioned earlier, the p-value of a two-sample t-test based on the data for non-diseased *X*_*k*_ and diseased *X*_*k*_ is represented as *P*_*jk*_. The fitted normal distribution based on the data for diseased *X*_*k*_ is denoted as *F*_*jk*_ (*x*). The marker score is calculated as

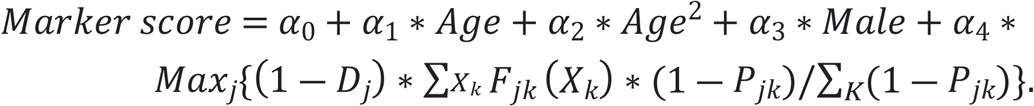

The regression coefficients are determined by fitting a logistic regression model, as outlined in Hosmer et al. [18]. The values of these coefficients for different transition probabilities are presented in Table 2.

**Table 2.**
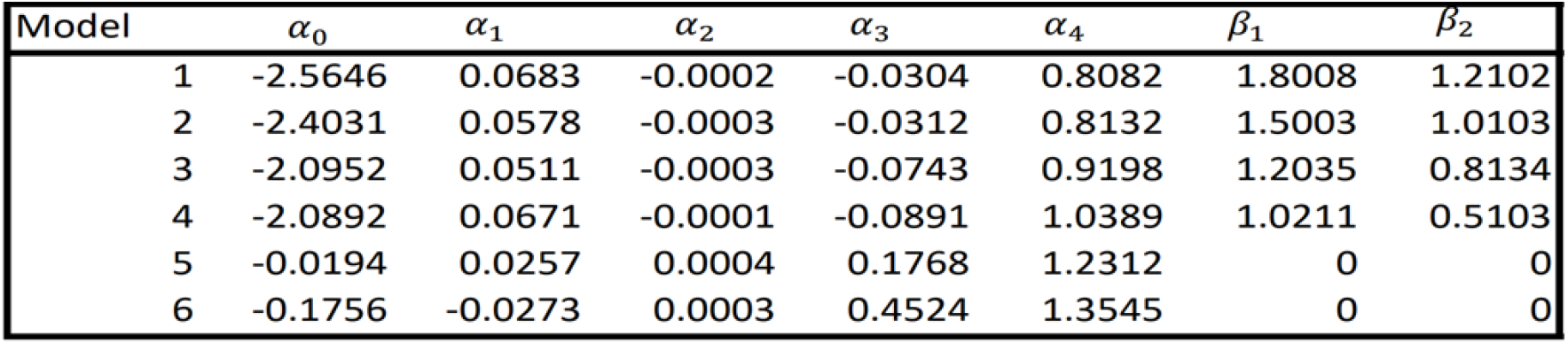
Transition probaility model coefficients.

Based on the models derived from steps 1 and 2, various interesting results can be obtained. For instance, individual-based 10-year risks for 15 diseases/symptoms can be estimated simultaneously by utilizing CL distribution probabilities as weights and the risks provided in Table 1. These predicted risks offer valuable information for precision health management. Another approach involves assigning different scores for different cluster levels and using the same method to define the health score, which is a weighted score employing cluster-level probabilities as weights. In this context, larger HS values indicate better health conditions. Moreover, when the HS exceeds 60, the participant is considered disease/symptom-free. A score between 45 and 60 suggests mild illness or the presence of one mild disease, such as hypertension or hyperlipidemia. Scores below 45 indicate the presence of at least two mild diseases or one severe disease, such as cancer. Essentially, the scores are assigned to construct a unique “disease map” for users, enabling them to gain insight into their own health conditions through the interpretation of their health score patterns. Subsequently, appropriate health management strategies can be implemented. For disease-free individuals, cutoffs for HS (based on age and gender) are identified, indicating that those below the cutoff have a significantly higher likelihood of developing diseases compared to those above it. Individuals satisfying the former condition are considered sub-healthy, while those meeting the latter criterion are classified as ordinary healthy. In the subsequent sections, we will compare the risks between ordinary healthy and sub-healthy individuals using data set A.

It should be noted that Table 1 exclusively displays the 10-year risks of different diseases and symptoms for individuals of all ages. However, we have also calculated 5 and 10-year risks for various age groups of interest, although the specific results are not reported here.

## 3. Results: Performance and validation

To validate the consistency and stability of the computation models, we compare the health scores generated by using Data B and two sub-data sets (Data B1 and Data B2) with the same assigned scores for cluster levels. Sub-data set B1 comprises 1,723,781 individuals collected between 2000 and 2009, observed for at least 10 years. Sub-data set B2 includes 228,847 individuals collected between 2003 and 2012, also observed for at least 10 years. We apply three computation models to Data B, Data B1, and Data B2, respectively, and compare the resulting health scores. To measure the difference between the models, we use the mean absolute percentage error (MAPE). Specifically, we calculate MAPE1, which represents the difference between the Data B-based model and the Data B1-based model, as well as MAPE2, which represents the difference between the Data B-based model and the Data B2-based model. The health scores for the current and future 10 years are computed for all models, and their 11-year MAPEs are compared. The computation of the health scores for the current and future years is identical, except that age is replaced with Age+t, while other health variables remain unchanged.

Table 3 presents the values of MAPE1 and MAPE2 for the current and future 10 years. The results indicate that both MAPE1 and MAPE2 are not only small but also very similar. This implies that the DKABio-HS is not only a consistent health score index system (with small MAPE values ranging from 1.22% to 1.52%) but also stable over time. The similarity between MAPE1 and MAPE2 is high, with the largest difference being only 0.11%.

**Table 3.** Comparison of MAPE1 and MAPE2.

| Year | MAPE1 | MAPE2 |
| --- | --- | --- |
| 0(current) | 1.44% | 1.52% |
| 1 | 1.41% | 1.49% |
| 2 | 1.38% | 1.47% |
| 3 | 1.35% | 1.44% |
| 4 | 1.33% | 1.42% |
| 5 | 1.31% | 1.40% |
| 6 | 1.29% | 1.38% |
| 7 | 1.27% | 1.37% |
| 8 | 1.25% | 1.35% |
| 9 | 1.23% | 1.34% |
| 10 | 1.22% | 1.33% |

Next, we proceed to compare the individual-based 10-year predicted disease risks with the true 10-year disease risks. We utilize Data B1 to develop the computation model and then apply this model to Data B2 to compute the predicted risks. The predicted risks and true risks based on Data B2 are compared in Table 4 for males and in Table 5 for females. In both tables, we group individuals into five levels using quintiles of the predicted risks as cutoffs for each disease/symptom. Within each level, we report the mean and standard deviation of the predicted risks as well as the true risks.

**Table 4.** Male 10-year predicted risks and true risks(%)*

|  | Level |  |  |  |  |  |  |  |  |  |
| --- | --- | --- | --- | --- | --- | --- | --- | --- | --- | --- |
|  | 1 |  | 2 |  | 3 |  | 4 |  | 5 |  |
| Disease/Symptom** | Predicted risk | True risk | Predicted risk | True risk | Predicted risk | True risk | Predicted risk | True risk | Predicted risk | True risk |
| Arthritis | 4.80(0.89) | 5.65 | 12.23(0.95) | 12.82 | 15.38(1.53) | 17.4 | 22.32(2.54) | 26.52 | 36.73(6.09) | 43.37 |
| Cancer | 0.21(0.04) | 0.21 | 0.40(0.13) | 0.39 | 1.32(0.46) | 1.21 | 3.55(0.98) | 2.90 | 11.38(4.22) | 8.99 |
| CER | 0.13(0.08) | 0.16 | 0.36(0.06) | 0.37 | 0.93(0.36) | 0.94 | 3.28(1.17) | 3.58 | 14.37(6.79) | 16.08 |
| CKD | 0.46(0.12) | 0.63 | 0.89(0.21) | 1.02 | 1.91(0.34) | 2.14 | 3.52(0.75) | 4.01 | 9.29(3.33) | 10.60 |
| COPD | 4.46(0.18) | 5.06 | 5.26(0.45) | 6.41 | 7.41(1.13) | 9.02 | 13.36(1.64) | 13.34 | 20.41(6.08) | 27.05 |
| DM | 0.18(0.09) | 0.2 | 0.56(0.25) | 0.59 | 2.82(1.24) | 3.08 | 7.82(1.68) | 9.08 | 13.93(2.42) | 16.15 |
| HD | 0.90(0.53) | 1.12 | 2.13(0.62) | 2.31 | 3.51(0.84) | 4.02 | 7.76(2.00) | 9.18 | 20.34(6.33) | 24.91 |
| HEPA | 0.95(1.11) | 1.47 | 5.01(1.32) | 5.98 | 9.50(0.95) | 11.56 | 10.72(0.09) | 13.26 | 12.64(2.08) | 15.27 |
| HL | 0.05(0.07) | 0.07 | 0.41(0.28) | 0.42 | 2.35(0.89) | 2.72 | 4.91(0.64) | 6.00 | 7.78(1.73) | 9.08 |
| HT | 0.20(0.24) | 0.24 | 1.43(0.67) | 1.50 | 6.48(2.51) | 6.91 | 16.69(3.60) | 19.24 | 32.24(5.71) | 37.52 |
| LC | 0.01(0.01) | 0.02 | 0.10(0.07) | 0.09 | 0.60(0.21) | 0.56 | 1.13(0.11) | 0.98 | 1.82(0.77) | 1.68 |
| PUB | 1.89(1.17) | 2.44 | 5.74(1.52) | 6.08 | 10.39(1.26) | 12.05 | 14.57(1.29) | 17.42 | 21.35(3.26) | 25.10 |
| SPY | 0.21(0.32) | 0.32 | 1.47(0.50) | 1.46 | 3.27(0.55) | 3.59 | 4.89(0.69) | 5.54 | 10.75(3.28) | 13.35 |
| PAIN | 1.68(0.15) | 1.87 | 3.26(0.55) | 2.95 | 4.23(0.59) | 6.26 | 7.40(1.39) | 8.63 | 19.07(5.85) | 22.80 |
| OMND | 0.18(0.01) | 0.21 | 0.19(0.01) | 0.20 | 0.25(0.03) | 0.26 | 0.48(0.13) | 0.53 | 4.92(4.57) | 5.58 |

\* Level predicted risk is the average of the predicted risks in the level; the number within the parentheses is the corresponding standard deviation.
\*\* CER: cerebrovascular disease; CKD: chronic kidney disease; COPD: chronic obstructive pulmonary disease; DM: diabetes mellitus; HD: heart disease; HEPA: hepatitis HL: hyperlipidemia; HT: hypertension; LC: liver cirrhosis; PUB: peptic ulcer and bleeding; SPY: somniphany; OMND: (old-age) major neurocognitive disorder

**Table 5.**
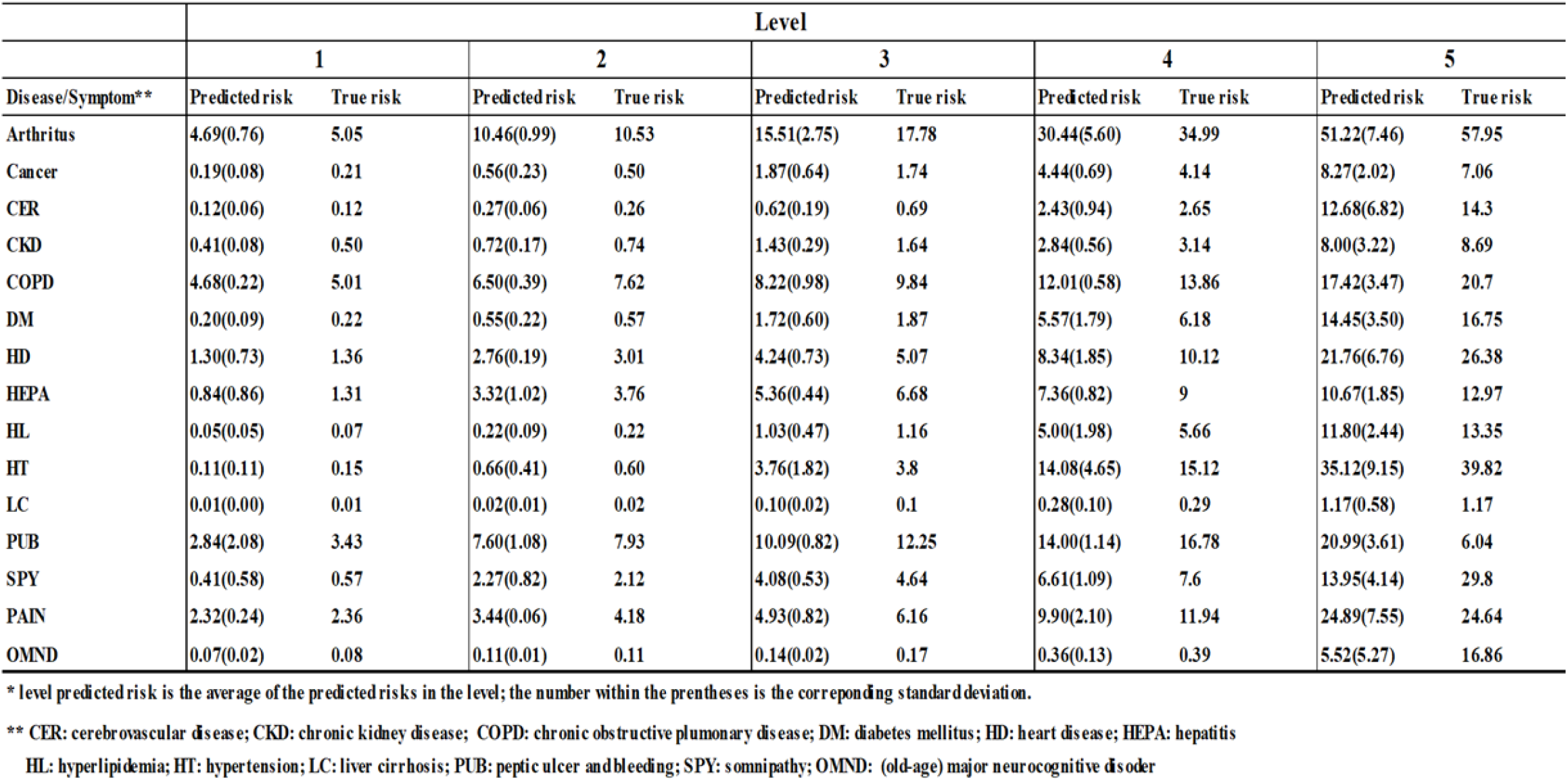
Female 10-year predicted risks and true risks(%)*

From Table 4 and Table 5, it becomes apparent that the true risk consistently increases as the risk level ascends for all diseases and symptoms. The differences between the predicted risks and true risks are generally small. However, in certain disease cases, particularly at higher risk levels, the differences tend to be relatively larger. This can be attributed to the presence of larger prediction variations in those cases. Nevertheless, we have found that the predicted risks and true risks are not statistically different in most instances. This indicates that our calculation of the predicted risk is reliable. Furthermore, we have observed considerable diversity in the risk differences between consecutive levels. For instance, in diseases like diabetes mellitus (DM), the risk differences between levels 3 and 4 (and 4 and 5) in Table 4 (and Table 5) are significantly greater than other differences. These findings highlight the importance of individuals exercising increased caution in managing their DM conditions when they reach risk level 3 or higher.

In the following analysis, we evaluate the performance of the “disease map” based on the application of Data A. As a reminder, health scores are categorized into four classes: M1 for individuals in an ordinary healthy status, M2 for individuals in a sub-healthy status, M3 for individuals with HS between 45 and 60, and M4 for individuals with HS below 45. Figures 1 (for males) and 2 (for females) present the risks of developing at least one new disease within t years (t=1,2,…,10) for individuals in classes M1 to M4.

**Figure 1.**
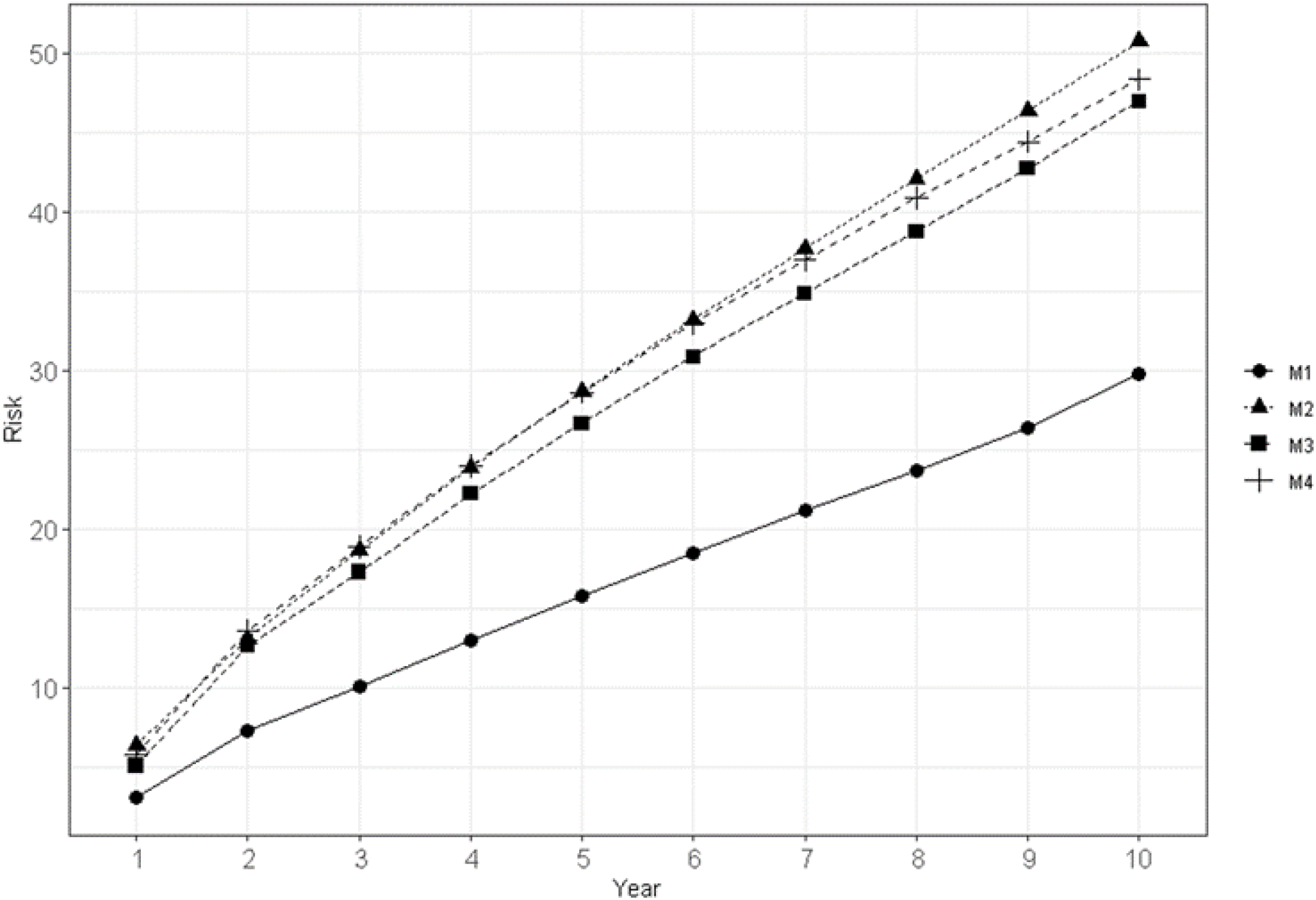
Disease risks(%) for male individuals in 4 classes ofhealth score

**Figure 2.**
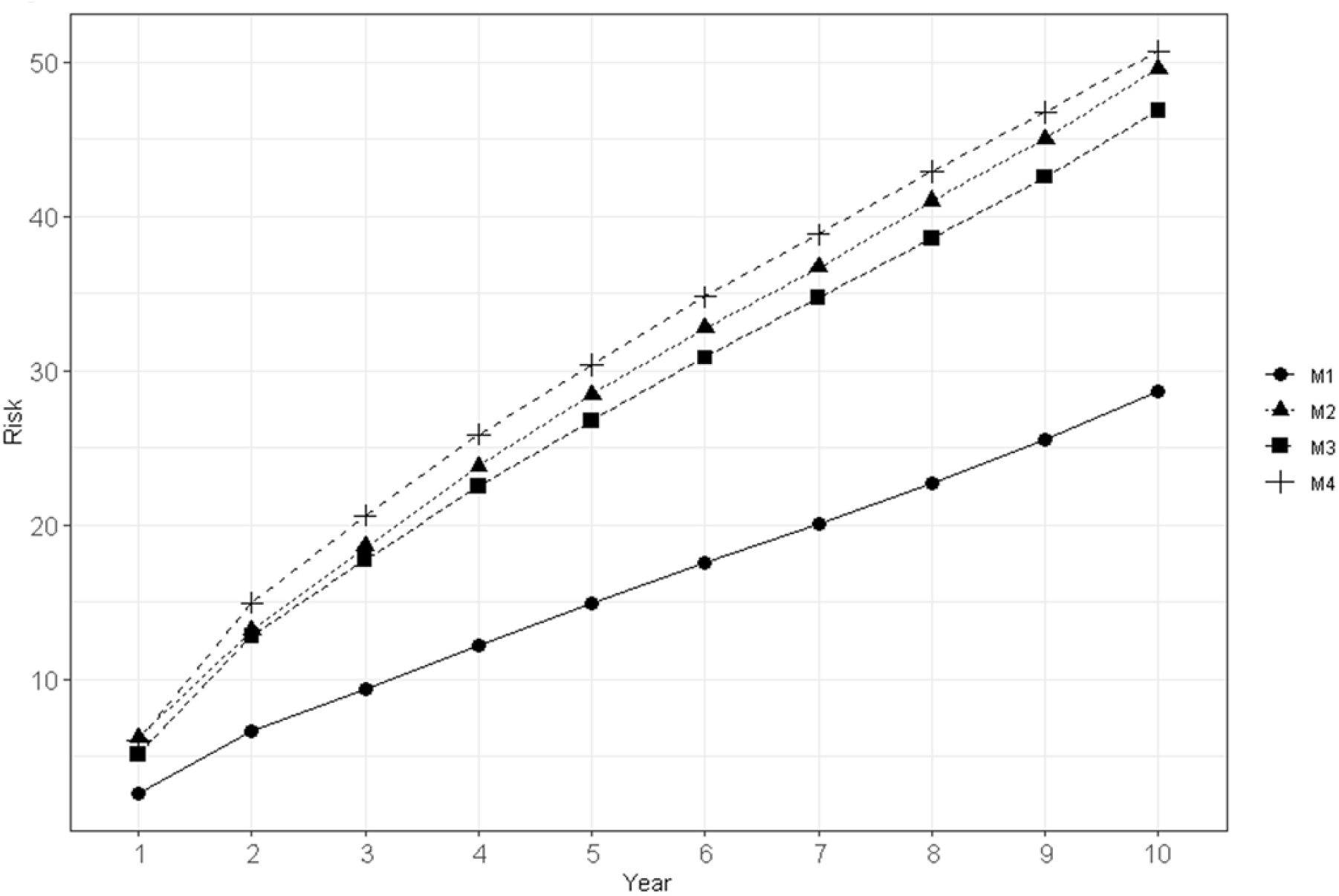
Disease risks(%) for female individuals in 4 classes of health score

We observe significant risk differences between class M1 (ordinary healthy individuals) and class M2 (sub-healthy individuals). The largest risk difference amounts to 21%. This result underscores the power of DKABio-HS in effectively distinguishing non-diseased individuals into more severe and less severe cases. If a non-diseased person is classified into M2, they should take their health conditions very seriously, considering more frequent health examinations or consultations with medical professionals.

In the male population, generally, M2 individuals exhibit the highest disease risk. Although the disease risks of M2 and M4 individuals appear indistinguishable in the first five years, with their largest difference being close to 0.5%, the difference increases to over 2.35% within 10 years. In contrast, the largest risk difference between M2 and M3 individuals is approximately 3.8%. Among the non-diseased groups (M2) and diseased groups (M3, M4), the M3 group has lower risk values within 10 years, while the M2 group has higher risk values. The overall disease risk ranking is M1, M3, M4, followed by M2. Although M2 individuals may seem more susceptible to diseases, the types of diseases that occur differ significantly among M1, M2, M3, and M4.

Table 6 highlights the top five diseases that occur in male individuals aged 65 and above within 5 and 10 years, across the M1-4 groups. Within 5 years, arthritis is the most frequent disease/symptom for M1 individuals, hypertension for M2 individuals, and heart disease for M3 and M4 individuals. Heart disease ranks second for M2 individuals and fourth for M1 individuals. Cancer does not feature in the top five diseases for M1 individuals, but it is the fifth most prevalent disease for M2 and M3 individuals and the fourth most prevalent for M4 individuals. Within 10 years, hypertension is the most frequently occurring disease/symptom for M1 and M2 individuals, and heart disease for M3 and M4 individuals. Heart disease ranks second for M2 individuals and third for M1 individuals. Cancer is the fifth most prevalent disease for M1, M2, and M3 individuals and the fourth most prevalent for M4 individuals.

**Table 6.** Top 5 diseases for male aged 65 and above*.

| Class | 0-5years |  |  |  |  | 0-10years |  |  |  |  |
| --- | --- | --- | --- | --- | --- | --- | --- | --- | --- | --- |
|  | 1 | 2 | 3 | 4 | 5 | 1 | 2 | 3 | 4 | 5 |
| M1 | ART | PUB | HT | HD | DM | HT | ART | HD | PUB | CAN |
| M2 | HT | HD | ART | DM | CAN | HT | HD | ART | DM | CAN |
| M3 | HD | HT | ART | DM | CAN | HD | HT | ART | DM | CAN |
| M4 | HD | ART | HL | CAN | DM | HD | HL | ART | CAN | DM |

\* ART: arthritis; CAN: cancer; DM: diabetes mellitus; HD: heart disease; HL: hyperlipid; HT: hypertension; PUB: peptic ulcer and bleeding.

In the female population, the performance of the disease map is similar to that of the male population, with some variations in the top five diseases. The M1 group still exhibits the lowest risk, and the largest risk difference between M1 and M2 is approximately 21%. However, the risk ranking among M2, M3, and M4 differs. M3 is ranked first, followed by M2 and then M4. Interestingly, in the female population, individuals in the M4 group appear to be more susceptible to diseases. Their highest risk of developing at least one disease within 10 years reaches 50.73%, although the risk difference between M2 and M4 is only about 2.16%.

Table 7 presents the top five diseases that occur in female individuals aged 65 and above within 5 and 10 years across the M1-4 groups. Within 5 years, arthritis is the most frequent disease/symptom for M1 individuals, hypertension for M2 individuals, and heart disease for M3 and M4 individuals. Heart disease ranks third for M1 and M2 individuals. Diabetes mellitus (DM) is the fifth most prevalent disease for M1 individuals and the fourth most prevalent for M1, M2, and M3 individuals. Within 10 years, hypertension remains the most frequently occurring disease/symptom for M1 and M2 individuals, while heart disease remains prevalent for M3 and M4 individuals. Heart disease ranks third for M1 and M2 individuals. DM is the fifth most prevalent disease for M1 individuals and the fourth most prevalent for M1, M2, and M3 individuals. Notably, cancer does not feature in the top five diseases for female M2, M3, and M4 individuals within the 0-10 year period.

**Table 7.** Top 5 diseases for female aged 65 and above*.

| Class | 0-5years |  |  |  |  | 0-10years |  |  |  |  |
| --- | --- | --- | --- | --- | --- | --- | --- | --- | --- | --- |
|  | 1 | 2 | 3 | 4 | 5 | 1 | 2 | 3 | 4 | 5 |
| M1 | ART | HT | HD | CAN | DM | HT | ART | HD | CAN | DM |
| M2 | ART | HT | HD | DM | CAN | HT | ART | HD | DM | HL |
| M3 | HD | ART | HT | DM | HL | HD | HT | ART | DM | HL |
| M4 | HD | HL | ART | DM | HT | HD | HL | ART | DM | HT |

\* ART: arthritis; CAN: cancer; DM: diabetes mellitus; HD: heart disease; HL: hyperlipid; HT: hypertension; PUB: peptic ulcer and bleeding.

## 4. Discussion

Health scores play a crucial role in capturing and measuring health and wellness, making the intangible aspects of health visible. Health scores are important in various directions of healthcare management. Firstly, they are valuable in interpreting data related to the outcomes of medical treatments or health management. By quantifying the illness or wellness of an individual, different health score ranges can be defined to represent various levels of health conditions. The DKABio disease map, for instance, provides this function not only for diseased individuals but also for non-diseased individuals. Secondly, a severity measure of illness like DKABio-HS, along with corresponding risk predictions, aids in identifying groups of patients with more severe illness, either currently or potentially, who may require additional treatment or care. Lastly, health scores can be highly useful in refining measures of healthcare resources at the individual or institutional level.

In this paper, we have proposed an unique AI system for measuring an individual’s health status and predicting 10-year risks for 15 diseases or conditions. Additionally, we have developed a disease map that allows for easy identification of disease severity using the health score. Notably, we have defined age-dependent sub-health conditions based on ranges of health scores and demonstrated that individuals meeting these conditions are more susceptible to diseases. To the best of our knowledge, this is the first formal definition of sub-health, which holds significant utility in precision health applications. We have demonstrated the consistency of HS, the efficiency of the disease map, and the accuracy of the risk predictions through the application of different databases. However, further external data verification is desirable to reinforce these findings.

The DKABio-HS and the derived risk predictions have been tested on large databases from distinct time periods and institutions in Taiwan. However, it is important to note that any health score, on its own, is not sufficient for comprehensive analyses required to assess healthcare outcomes and treatment effectiveness. It is crucial to use the health score in conjunction with other analytic tools that measure other aspects of care. For example, an analytic tool that provides recommendations for potential disease prevention in individuals would be a valuable addition for care providers or users.

## Data Availability

We understand that the data sets used in the manuscript cannot be shared due to the data purchase contract agreement. We will make a note of this in the manuscript and acknowledge that the data sets can be purchased from Mei Jau Health Management Institution and the National Health Insurance, Taiwan

## Acknowledgement

This research was partially supported by the National Science and Technology of Taiwan and Taipei Medical University.

## Statement on Conflicts of interest

No

| Summary Table |  |
| --- | --- |
| Section | Key Points |
| **Objective** | Accurate morbidity estimation using DKABio -clusters and HS health score |
| **Precision Health** | Focus on proactive disease prevention using AI and data science |
| **Health Score (HS)** | Numerical value indicating health status; higher score = better health |
| **Disease Map** | Classification into healthy, sub-healthy, and diseased states for tailored health management |
| **Data Sources** | NHIRD (2 million participants) and Mei Jau database (0.75 million participants) |
| **Participants** | 750,000 individuals aged 20+ with comprehensive health data |
| **Methodology** | Hierarchical clustering and logistic regression on 148 health variables |
| **Clustering Algorithm** | Partitions participants into clusters based on disease status and health variables |
| **Comorbidity Score** | Derived from Charlson Comorbidity Index for 15 chronic diseases |
| **Age Indexes** | Calculated using specific formulas for males and females |
| **Transition Probability** | Estimates health score distribution and disease risk transitions |
| **Validation** | High stability with MAPEs below 0.1%, minimal variations between datasets |
| **Predictive Accuracy** | Close match between predicted and true 10-year disease risks |
| **10-Year Disease Risks** | Provided for 15 diseases/conditions based on cluster levels |
| **Health Score Interpretation** | Scores above 60: disease-free; below 45: significant health issues |
| **Health Management** | Identifies sub-healthy individuals for preventive action |
| **Ethical Approval** | Approved by NTU Ethical Review Committee (NTU-REC No.: 202402EM002) |
| **Impact** | Enhances precision health management through AI and data science |
| **Future Directions** | Further research, broader application in healthcare settings |

